# Comorbidity analysis and clustering of endometriosis patients using electronic health records

**DOI:** 10.1101/2025.02.13.25322244

**Authors:** Umair Khan, Tomiko T. Oskotsky, Bahar D. Yilmaz, Jacquelyn Roger, Ketrin Gjoni, Juan C. Irwin, Jessica Opoku-Anane, Noémie Elhadad, Linda C. Giudice, Marina Sirota

**Affiliations:** Bakar Computational Health Sciences Institute, University of California, San Francisco, San Francisco, CA; Biological and Medical Informatics Graduate Program, University of California, San Francisco, San Francisco, CA; Department of Obstetrics, Gynecology, and Reproductive Sciences, Center for Reproductive Sciences, University of California, San Francisco, San Francisco, CA; Pharmaceutical Sciences and Pharmacogenomics Graduate Program, University of California, San Francisco, San Francisco, CA; Robert Wood Johnson Medical School, Rutgers University, New Brunswick, NJ; Department of Biomedical Informatics, Columbia University, New York, NY

**Keywords:** endometriosis, electronic health records, comorbidities, unsupervised clustering

## Abstract

Endometriosis is a prevalent, complex, inflammatory condition associated with a diverse range of symptoms and comorbidities. Despite its substantial burden on patients, population-level studies that explore its comorbid patterns and heterogeneity are limited. In this retrospective case-control study, we analyzed comorbidities from over forty thousand endometriosis patients across six University of California medical centers using de-identified electronic health record (EHR) data. We found hundreds of conditions significantly associated with endometriosis, including genitourinary disorders, neoplasms, and autoimmune diseases, with strong replication across datasets. Clustering analyses identified patient subpopulations with distinct comorbidity patterns, including psychiatric and autoimmune conditions. This study provides a comprehensive analysis of endometriosis comorbidities and highlights the heterogeneity within the patient population. Our findings demonstrate the utility of EHR data in uncovering clinically meaningful patterns and suggest pathways for personalized disease management and future research on biological mechanisms underlying endometriosis.

## Introduction

Endometriosis is a chronic and often debilitating inflammatory and systemic condition that affects millions of individuals worldwide.^1^ It is characterized by the presence of endometrial-like tissue outside the uterus, leading to inflammation, scarring, and adhesions.^2^ Endometriosis is estimated to affect approximately 10% of reproductive-aged individuals with a uterus, making it a significant public health concern. The condition is associated with a wide range of symptoms, including chronic pelvic pain, infertility, dysmenorrhea, and gastrointestinal disorders, all of which contribute to a substantial burden on patients’ quality of life.^3^

Despite its prevalence, diagnosing and managing endometriosis remains challenging due in part to wide variability in manifestation. Patients often take years to reach a diagnosis, during which symptoms are frequently misattributed to other conditions and disease can progress.^4^ Even though many clinicians use clinical judgment to diagnose and initiate treatment, the diagnostic delay is often further influenced by the reliance on surgery for definitive diagnosis. Furthermore, treatment options are complex and response rates are variable. While hormonal therapies and surgical interventions can provide symptom relief, they are associated with side effects and a high likelihood of symptom recurrence.^5^ Beyond these clinical hurdles, endometriosis imposes a substantial psychosocial burden on patients, who may face stigma, invalidation of their pain, and diminished quality of life.^6^

Although many smaller studies have provided valuable insights into endometriosis’ heterogeneity^7–10^, they often cannot capture broad population-level features that characterize the disease. Electronic health records (EHRs) offer an opportunity to study large patient populations and uncover patterns that may not be apparent in smaller-scale studies.^11^ By leveraging this data, researchers can identify trends and associations that might otherwise go unnoticed. The application of EHRs to study endometriosis is particularly promising, as it allows for a more comprehensive examination of the disease and its comorbidities in diverse and extensive patient populations.

Prior studies investigating endometriosis using EHRs have highlighted their potential for providing insights into the condition. For instance, Choi et al. analyzed patient information from South Korea’s Health Insurance Review and Assessment data and found 44 ICD-10 codes that were significantly associated with endometriosis.^12^ Also, Estes et al. examined administrative health claims data from Optum’s Clinformatics DataMart to compare the incidence of mental health outcomes in patients in the United States with and without endometriosis, finding an increased risk for anxiety, depression, and self-directed violence.^13^ However, these studies focus on specific aspects of the disease, such as particular classes of comorbidities or certain patient subpopulations, and do not attempt to validate their findings across independent data sources. Along similar lines, Urteaga et al. used patient-reported data from a smartphone app to identify subtypes of endometriosis.^14^ Though not built around structured EHR data, this work demonstrates the utility of data-driven methods in characterizing this complex disease.

In this work, we aim to address these gaps by analyzing the comorbidities of endometriosis patients across multiple medical centers. Specifically, we examine data from the University of California, San Francisco (UCSF), and five other UC medical centers, utilizing odds ratio analysis and unsupervised clustering techniques to identify and characterize patterns of diagnoses associated with endometriosis (Figure 1a). Our analysis compares endometriosis patients with matched controls, considering both pre-endometriosis conditions and diagnoses across the full patient timeline. Clustering is then performed on the endometriosis patients to identify subgroups that may reflect different disease trajectories or phenotypes. This dual focus enables us to capture the overall landscape of endometriosis comorbidities compared to controls, while also examining the heterogeneity within the endometriosis patient population. By uncovering these patterns, we aim to contribute to a more comprehensive understanding of endometriosis, its clinical features, and its impact on patient health.

**Figure 1.**
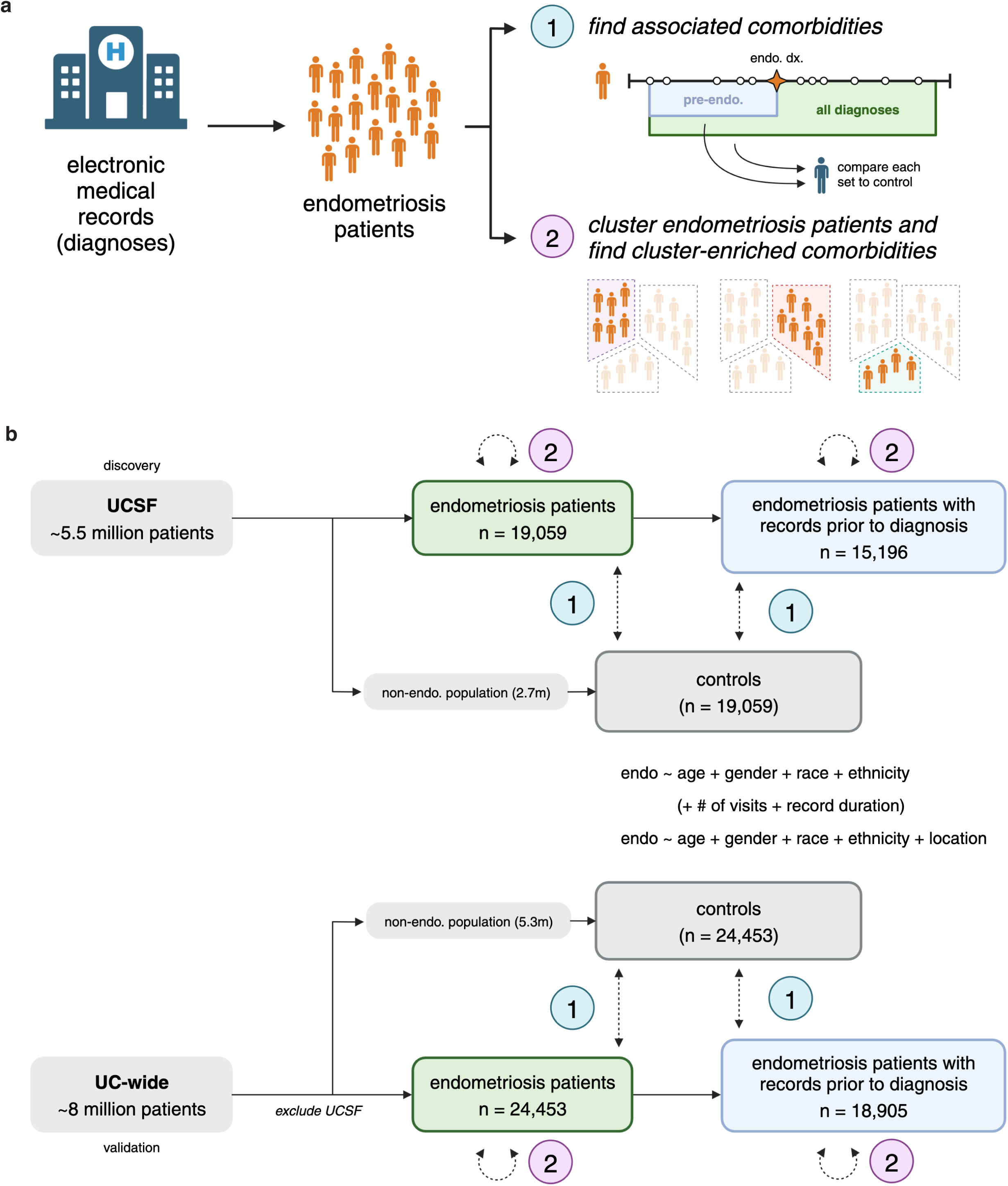
Study overview. (a) Overview of study design. Endometriosis patients were selected from electronic medical records by diagnoses. We then (1) compute odds ratios for comorbidities by comparing against matched controls, and (2) identify groupings of endometriosis patients using unsupervised clustering. (b) Details of patient selection process. We select cases using the “endometriosis” concepts from the OMOP concept hierarchy, and match against controls from the non-endometriosis population using propensity score matching on both demographic and healthcare utilization covariates.

## Methods

### Patient selection and timeline definition

Patients were selected from two separate de-identified EHR databases, both of which conform to the Observational Medical Outcomes Partnership^15^ (OMOP) Common Data Model^16^ (CDM) schema. We initially performed our analysis using UCSF’s records, which run from 1988 and comprise over 5.5 million patients. We then expanded our scope to the University of California’s Health Data Warehouse (UCHDW), which aggregates medical records from over 8 million patients seen at six University of California medical centers, spanning 2012 to the present day. We explicitly excluded UCSF patients from our UCHDW queries to avoid patient overlap (Figure 1b).

We defined endometriosis cases as patients who at any point in their medical history were assigned at least one of the 49 standard Systematized Nomenclature of Medicine (SNOMED) condition IDs descended from “endometriosis” in OMOP’s concept hierarchy (Supplementary Table 1). Conversely, we identified a background population by selecting patients who have at least one condition and were never diagnosed with endometriosis. We selected controls via 1:1 propensity score matching against cases on age, gender, race, and ethnicity (and location for UCHDW patients) using the MatchIt^17^ package in R (version 4.5.5, nearest neighbor method) (Supplementary Figure 1). We also selected groups of healthcare utilization matched controls by adding number of visits and record duration as additional covariates for matching. These utilization-matched controls were used to filter significant associations, as explained in the following sections.

Our initial analysis considers all diagnoses across any given patient’s record. To examine the pattern of comorbidities observed prior to endometriosis, we repeat the following methods using only patients with conditions assigned prior to their first endometriosis diagnosis, and their matched controls. We restrict the condition set to those that appear in the record before the first endometriosis condition, or conditions assigned at any time to the remaining matched controls.

### Association analysis

For each of the SNOMED conditions ever assigned to a patient, we calculated the odds ratio between cases and controls and computed a p-value by the hypergeometric test (or Fisher’s exact test if any counts were less than five). To determine significantly enriched conditions, we compared the p-values against a Bonferroni-corrected threshold with = 0.05.

To ensure our significantly enriched conditions are robust to variation in the selected control patients, we repeat this process across thirty separately selected control groups. We then aggregate results by computing the mean odds ratio and a new p-value, defined as twice the mean p-value across the thirty control iterations.^18^

In addition, we account for healthcare utilization in our analysis by repeating the procedure using control groups matched on utilization covariates, as described in the previous section. A condition is considered significant if it is first identified in the non-utilization-matched comparison and remains significant after adjusting for healthcare utilization. This approach eliminates conditions that appear significant solely due to increased healthcare interactions, while minimizing false protective associations from control patients with higher overall healthcare needs.

### Clustering and cluster enrichment

To identify clusters of endometriosis patients, we first construct a matrix where each row represents an endometriosis patient, and each column represents a condition. An entry is set to 1 if the corresponding patient has the given condition, and 0 otherwise. To identify clusters, we applied principal component analysis to this matrix to reduce each patient’s representation down to 1,000 dimensions, explaining ∼80% of the variance. We then constructed a neighborhood graph and applied the Leiden community detection algorithm^19^ using scanpy^20^. To examine each of the resulting clusters, we perform the odds ratio analysis procedure described previously, with the “cases” being endometriosis patients in the cluster under consideration and the “controls” being patients in all other clusters. We then identify significantly enriched conditions that are exclusive to each cluster.

### Comparing data sources

We compared both the association and clustering analyses across the two data sources. For the association analysis, we first found the intersection of significantly enriched conditions across the two data sources and then computed the Pearson correlation coefficient and associated p-value between the log-odds ratios of those conditions. For the clustering analysis, we matched each cluster found at UCSF to the UCHDW cluster with the most overlapping significant conditions (Figure 3a), if there was one, and visualized the results as a bipartite graph. Note that only clusters that have overlapping conditions between the two data sources are visualized in the graphs.

**Figure 2.**
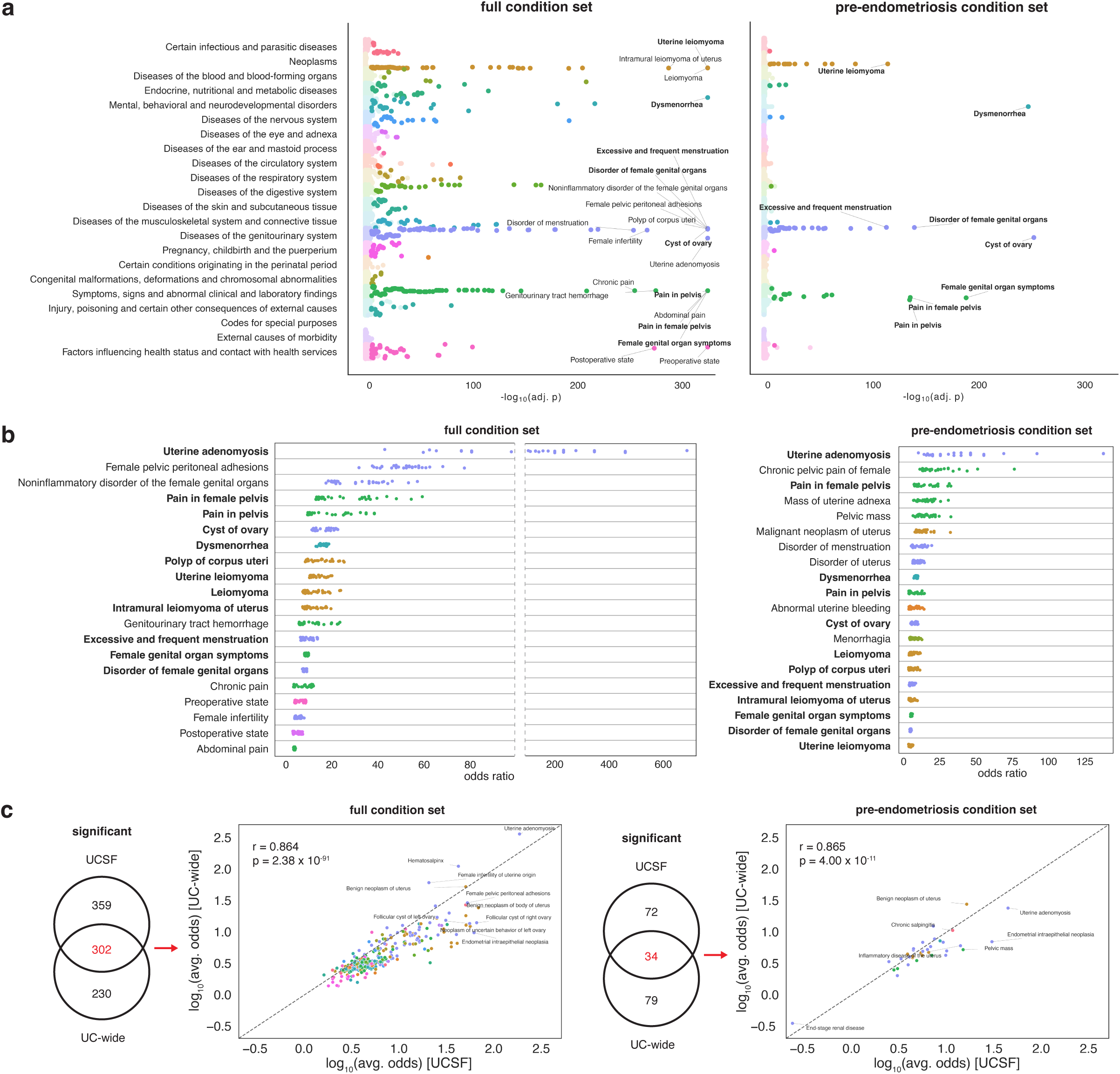
Comorbidity analysis and validation. (a) Manhattan plots of comorbidities observed at UCSF, organized by ICD chapter. (b) Strip plots of top twenty comorbidities (by aggregate p-value) observed at UCSF, showing computed odds ratios across thirty replicate control groups. (c) Comparison to UC-wide analysis, showing number of significant overlapping comorbidities and concordance of odds ratios for overlapping conditions.

**Figure 3.**
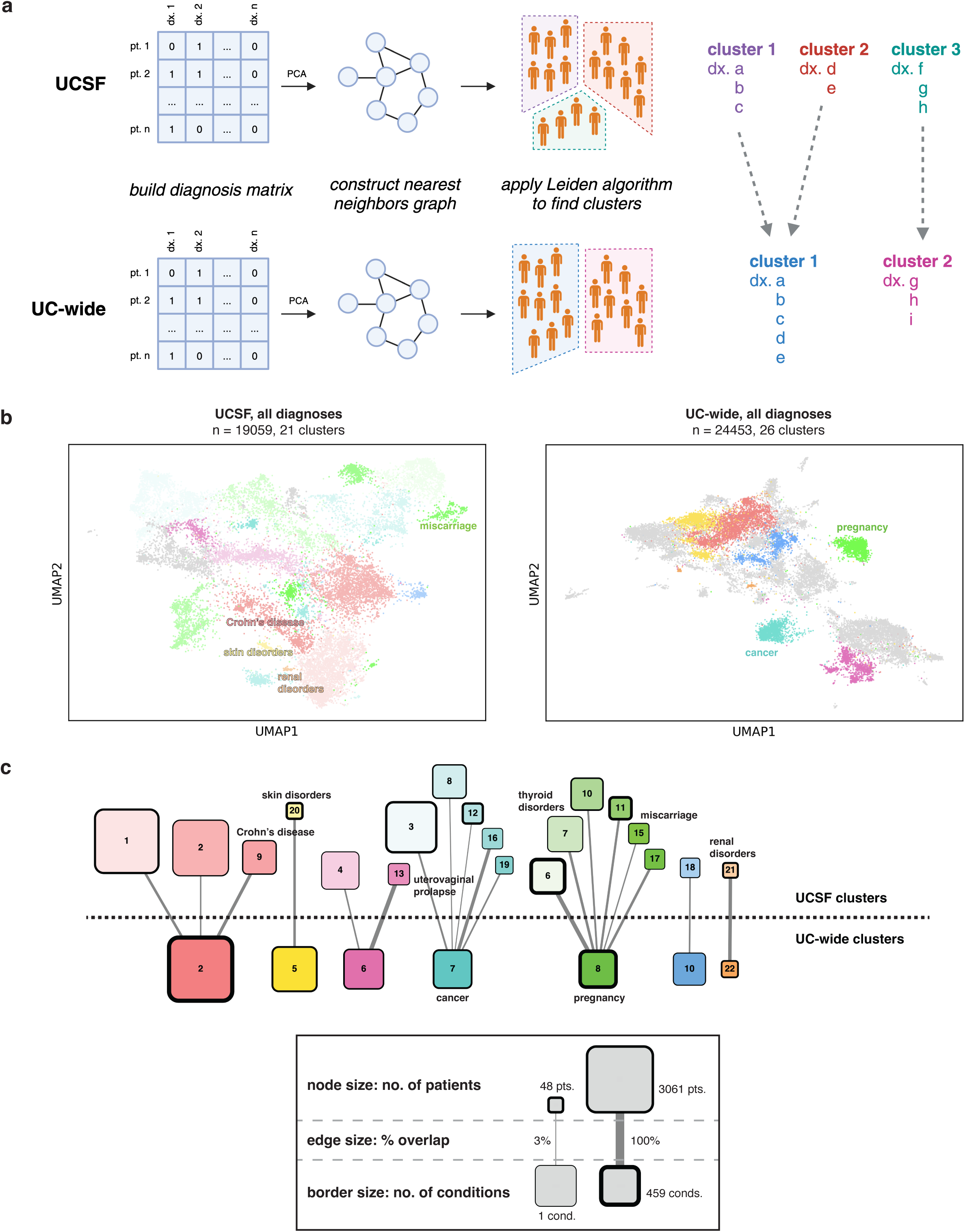
Clustering overview and results, all conditions. (a) Overview of clustering process. From the initial matrix of diagnoses, a nearest neighbors graph is constructed based on patient-to-patient distances. The Leiden algorithm is then applied to this graph. Each cluster from UCSF is matched to the UC-wide cluster with the greatest proportion of overlapping significantly enriched conditions. (b) Visualization of clusters computed using all diagnoses overlaid on two-dimensional UMAP representations of endometriosis patients, at UCSF and UC-wide. (c) Concordance between UCSF and UC-wide clusters. Node and edge properties map to cluster characteristics as specified in the legend.

### Visualizations

We applied the Uniform Manifold Approximation and Projection (UMAP) algorithm^21^ to the diagnosis matrix like the one used for the clustering analysis but including all endometriosis patients and their closest matched controls, to visualize the comorbidity structure. We used Cytoscape^22^ to construct the bipartite graph visualizations and associated legends for the clustering comparison between the two data sources. All other illustrations were created with BioRender, and all other data visualizations were generated using matplotlib.^23^ To annotate SNOMED concepts with ICD chapter labels for visualization, we used the SNOMED CT to ICD-10-CM Map provided in the Unified Medical Language System created by the National Library of Medicine.^24^

## Results

### Patient characteristics

We identified a total of 19,059 endometriosis patients at UCSF and an additional 24,453 endometriosis patients across UCHDW (Table 1). These patients had a mean age of 52.6 years at UCSF and 46.5 years in the UCHDW. In both data sources, endometriosis patients were predominantly white (51.7% at UCSF, 53.9% in the UCHDW) with “Asian” and “other” being the next two most common classifications. Similarly, most patients (76.1% at UCSF, 71.8% in the UCHDW) were identified as not Hispanic or Latino. Most of our UCHDW patients come from records at two large centers (44.6% and 24.8%, respectively) with the remaining being split across the three other centers.

**Table 1.** Patient characteristics at UCSF and across the other UC medical centers analyzed.

|  | **UCSF** | **UC-wide** |
| --- | --- | --- |
| **total (n)** | 19059 | 24453 |
| **age (mean, sd)** | 52.6 (sd 14.3) | 46.5 (sd 11.9) |
| **gender** |  |  |
| female | 18995 (99.6%) | 24411 (99.8%) |
| male | 36 (0.2%) | 34 (0.1%) |
| unknown | 21 (0.1%) | 6 (0.0%) |
| other | 7 (0.0%) | 2 (0.0%) |
| **race** |  |  |
| white | 9859 (51.7%) | 13168 (53.9%) |
| Asian | 2980 (15.6%) | 2767 (11.3%) |
| other | 2327 (12.2%) | 3170 (13.0%) |
| unknown | 2150 (11.3%) | 2628 (10.7%) |
| Black or African American | 1105 (5.8%) | 1404 (5.7%) |
| Native Hawaiian or Other Pacific Islander | 491 (2.6%) | 121 (0.5%) |
| American Indian or Alaska Native | 147 (0.8%) | 112 (0.4%) |
| multirace | - | 1083 (4.4%) |
| **ethnicity** |  |  |
| Not Hispanic or Latino | 14502 (76.1%) | 17560 (71.8%) |
| unknown | 2570 (13.5%) | 2378 (9.7%) |
| Hispanic or Latino | 1987 (10.4%) | 4515 (18.5%) |
| **location** |  |  |
| UC center #1 | - | 10901 (44.6%) |
| UC center #2 | - | 6069 (24.8%) |
| UC center #3 | - | 3638 (14.9%) |
| UC center #4 | - | 2986 (12.2%) |
| UC center #5 | - | 859 (3.5%) |

### Association analysis across the full condition set identifies diverse range of comorbidities, consistent across multiple medical centers

When considering the full condition set, our analysis revealed 661 significantly enriched (aggregated adjusted p-value < 0.05) comorbidities at UCSF spanning across nearly all ICD chapters (Figure 2a), reflecting the diverse clinical presentations associated with endometriosis (Supplementary Table 2). Among the most significantly enriched conditions were uterine adenomyosis (OR = 181), pelvic peritoneal adhesions (OR = 51.1), non-inflammatory disorders of the female genital organs (OR = 30.2), pain in female pelvis (OR = 26.3), and cyst of ovary (OR = 16), all of which were also significant in the UCHDW (Figure 2b). We also see significant enrichments for female infertility (OR = 5) and general autoimmune disease (OR = 4.3), consistent with prior literature on endometriosis-associated infertility^25^ and systemic inflammation^26^. Interestingly, we identified several conditions less commonly reported in smaller studies, including migraines^27–29^ (OR = 4), gastroesophageal reflux disease^30^ (OR = 3.6), asthma^31^ (OR = 2.5), and vitamin D deficiency^32^ (OR = 3.8). We found that 302 conditions of these conditions (45% of the complete set) were significantly enriched in the UCHDW data as well (Supplementary Table 3), with statistically significant correlation of the log-odds ratios (Pearson r = 0.864 and p = 2.38 x 10^-91) (Figure 2c).

### Association analysis across the pre-endometriosis condition set emphasizes genitourinary conditions

When considering the pre-endometriosis condition set, our analysis revealed 106 significantly enriched (aggregated adjusted p-value < 0.05) comorbidities at UCSF (Supplementary Table 4). These conditions span a narrower range of ICD chapters, with most enriched conditions falling into those spanning the genitourinary system, related symptoms, and neoplasms (Figure 2a). Among the most significantly enriched conditions were cyst of ovary (OR = 6.6), dysmenorrhea (OR = 8.3), female genital organ symptoms (OR = 4.9), disorder of female genital organs (OR = 4.3), and pain in female pelvis (OR = 15.2) (Figure 2b). We also see a significant enrichment for increased cancer antigen 125 (OR = 17.9), consistent with prior literature describing the relationship between this biomarker and endometriosis.^33^ Interestingly, the association between migraines and endometriosis remained significant even prior to the diagnosis of endometriosis (OR = 2). We found that 34 of these conditions (32% of the complete set) were significantly enriched in the UCHDW data as well (Supplementary Table 5), with statistically significant correlation of the log-odds ratios (Pearson r = 0.865 and p = 4.00 x 10^-11) (Figure 2c). The pre-endometriosis analysis in the UCHDW dataset found several notable negative associations, with odds ratios less than one, including hyperlipidemia (OR = 0.67) and mixed hyperlipidemia (OR = 0.67).

### Clustering analysis identifies endometriosis patient subpopulations

Our unsupervised clustering analysis identified distinct subpopulations of endometriosis patients, characterized by shared diagnosis patterns. Specifically, we identified 21 clusters in the UCSF cohort and 26 clusters in the UCHDW cohort when analyzing the full condition set (Figure 3b). Similarly, the pre-endometriosis condition set yielded 31 clusters at UCSF and 41 clusters in the UCHDW cohort (Figure 4a). These clusters revealed diverse patterns of comorbidities, with some clusters being dominated by autoimmune disorders, others by pregnancy complications, and still others by psychiatric conditions. Annotation of clusters suggested biologically and clinically meaningful subgroupings, which may reflect underlying heterogeneity in the disease’s pathophysiology or healthcare utilization patterns. Comparing cluster concordance across our two independent datasets, we saw UC-wide clusters highlighted by pregnancy and cancer-related conditions when considering all diagnoses (Figure 3c), and pregnancy and urinary tract conditions when considering pre-endometriosis diagnoses (Figure 4b). Clusters at UCSF also highlighted a number of other disease categories, including skin disorders, renal disorders, and mental health conditions.

**Figure 4.**
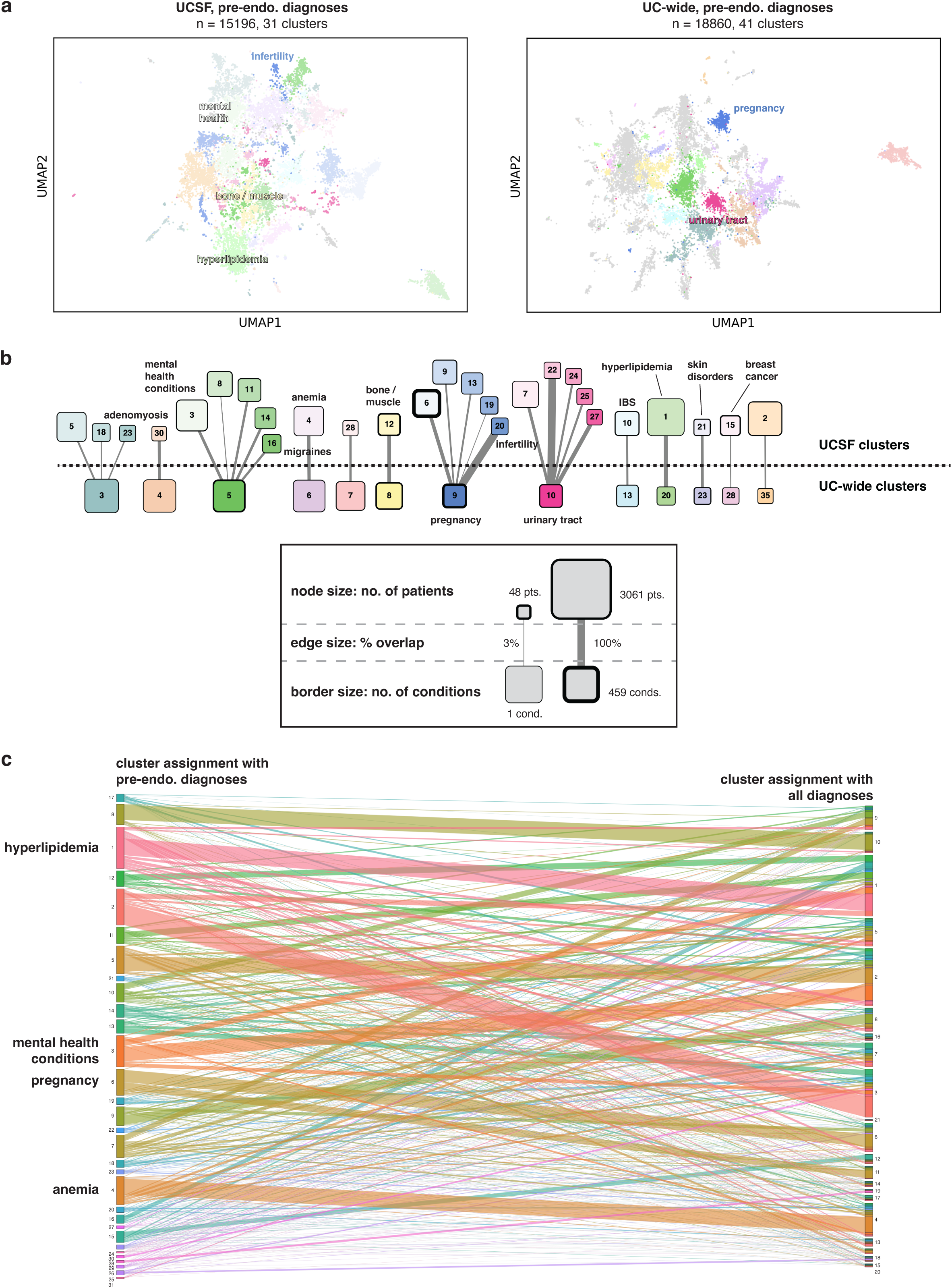
Clustering results, pre-endometriosis conditions. (a) Visualization of clusters computed using pre-endometriosis diagnoses overlaid on two-dimensional UMAP representations of endometriosis patients, at UCSF and UC-wide. (b) Concordance between UCSF and UC-wide clusters. Node and edge properties map to cluster characteristics as specified in the legend. (c) Alluvial plot showing change in cluster membership when considering pre-endometriosis and all diagnoses at UCSF.

In addition to a strong concordance of associations when comparing pre-endometriosis diagnoses to all diagnoses (Supplementary Figure 3), analysis of the patient clusters at UCSF reveals that certain groups of patients remain consistently clustered across both condition sets (Figure 4c). This suggests that these groups of patients, initially assigned to pre-endometriosis clusters associated with hyperlipidemia, mental health conditions, pregnancy, and anemia, might experience similar clinical trajectories after their endometriosis diagnosis.

## Discussion

Our analysis of comorbidities in endometriosis patients highlights findings that align closely with previous investigations. Many of the observed associations between endometriosis and its comorbidities, such as chronic pain conditions^34^ and gastrointestinal diseases^35^, are consistent with established literature. In addition, our analysis identified several less-frequently reported associations, demonstrating the power of this data-driven approach. For instance, the protective associations with hyperlipidemia and mixed hyperlipidemia in the UCHDW dataset are particularly interesting in the context of a body of literature identifying statins as potential therapeutic avenues for endometriosis^36–39^, since patients with these conditions are likely taking statins as treatment. Similarly, the repeated appearance of migraines as a significant association, both before and after endometriosis diagnosis, underscores the emerging concept of repurposing drugs from comorbidities to treat endometriosis pain – in a recent study, Fattori et al. showed that a migraine drug decreased disease burden and pain in a mouse model of endometriosis.^40^ These results provide further evidence of the broad and multifaceted clinical impact of endometriosis, underscoring the importance of a comprehensive approach to its management.

Notably, many of these associations replicated across two separate data sources and time periods, reinforcing the robustness of our findings. This replication suggests that the observed patterns are not artifacts of a single dataset or limited to a specific population but instead represent generalizable trends across diverse healthcare environments. Such consistency enhances the credibility of our results and their relevance to broader patient populations.

Through unsupervised clustering, we identified distinct subpopulations of endometriosis patients characterized by unique patterns of comorbidities. These clusters reveal novel insights into the heterogeneity of endometriosis, suggesting that patients may fall into clinically meaningful subgroups based on their associated diagnoses. Importantly, some of these subpopulations also replicated across different datasets and time periods, further supporting the validity of our approach. These findings may serve as a foundation for future studies aiming to tailor treatments or management strategies to specific patient subgroups. For instance, these subgroups may be associated with differing patterns of drug utilization, providing a starting point for examining patient outcomes.

Our study has several strengths worth highlighting. First, the use of data from multiple medical centers and time periods allows for a more comprehensive and generalizable analysis of endometriosis and its comorbidities. Second, the robust control replication method employed in our analyses mitigates the risk of false-positive associations, which is often a concern with studies of this nature. Finally, our use of EHR data provides a valuable perspective on real-world population patterns, which can inform clinical practice and policy.

### Limitations of the study

As with any research leveraging EHRs, our analysis is subject to inherent data issues, including missingness, patients moving between healthcare systems, and coding differences across institutions. Additionally, our selection criteria were deliberately permissive, as we defined endometriosis cases based on EHR-documented diagnoses rather than surgically confirmed cases. While this approach maximized the size and diversity of our sample, it may have introduced misclassification bias. Moreover, the case-control design of our study precludes any conclusions about causality or temporality in the observed associations. Our analysis is also geographically constrained, being limited to medical centers in California, which may limit the applicability of our findings to other regions or populations. In particular, populations with limited access to care or inadequate insurance coverage may experience very different outcomes.

Despite these limitations, our study provides valuable insights into the comorbidities and heterogeneity of endometriosis. By leveraging data from multiple institutions and time periods, employing robust statistical analysis methods, and considering the impact of healthcare utilization patterns on EHR data, we address several of the challenges inherent to EHR-based research. Our findings contribute to a growing body of evidence that underscores the complexity of endometriosis and highlights the potential for EHR data to advance our understanding of this condition on a population level.

### Conclusions

Our study provides a comprehensive analysis of endometriosis comorbidities using large-scale EHR data from multiple medical centers. By comparing endometriosis patients to matched controls, we captured both the broad range of associated comorbidities and the specific patterns preceding diagnosis. Importantly, our clustering analysis revealed meaningful subgroups within the endometriosis patient population, highlighting the heterogeneity of the condition and suggesting potential pathways for personalized management strategies.

The replication of key findings across two independent datasets reinforces the robustness and generalizability of our results. This consistency underscores the value of leveraging EHR data to study complex, heterogeneous conditions like endometriosis at scale. However, our findings also emphasize the challenges inherent in analyzing real-world data, such as biases introduced by healthcare utilization patterns and limitations in diagnostic coding.

Looking ahead, our work lays a foundation for future studies to explore further relationships between endometriosis and its comorbidities, as well as the biological mechanisms underlying the observed heterogeneity. Integrating genomic, clinical, and patient-reported data with EHR-based findings may further enhance our understanding and ultimately support the development of targeted diagnostic tools and treatment strategies, including with novel machine learning approaches. By advancing knowledge of endometriosis and its comorbidities, this research contributes to ongoing efforts to improve patient care, reduce diagnostic delays, and address the significant burden of this disease.

## Resource availability

### Lead contact

Requesters for further information and resources should be directed to and will be fulfilled by the lead contact, Marina Sirota.

### Materials availability

This study did not generate any new materials.

### Data and code availability

The data that support the findings of this study are not openly available to individuals unaffiliated with UCSF due to the sensitivity of medical records, with the exception of collaborators. Individuals not affiliated with UCSF may set up an official collaboration with a UCSF-affiliated investigator by reaching out to the lead contact, Marina Sirota. UCSF-affiliated individuals may contact UCSF’s Clinical and Translational Science Institute or the UCSF’s Information Commons team for more information. UC-wide data is only available to UC researchers who have completed analyses in their respective UC first and have provided justification for scaling their analyses across UC health centers. Censored code for the analysis and visualizations in this study can be found at https://github.com/khanu263/comorbidities-clustering-endo.

## Acknowledgements

The authors would like to thank Parker Grosjean, Duncan Muir, Chimno Nnadi, Michael Keiser, Stacey Missmer, Christina Theodoris, Tony Capra, and all members of the Sirota Lab for insightful conversations and guidance throughout the preparation of this work. This manuscript was supported by the Eunice Kennedy Shriver National Institute for Child Health and Human Development, P01HD106414 (UK, TTO, JCI, JO, LCG, MS), and the National Institute of General Medical Sciences, T32GM067547 (UK) and T32GM142516 (KG).

## Author contributions

Conceptualization, UK, TTO, KG, LCG, and MS; Methodology, UK, JR, and MS; Software, UK; Investigation, UK and TTO; Writing – Original Draft, UK, TTO, and BDY; Writing – Review & Editing, all authors; Supervision, JCI, JO, NE, LCG, and MS; Project Administration, MS; Funding Acquisition, LCG and MS.

## Declaration of generative AI and AI-assisted technologies in the writing process

During the preparation of this work the authors used ChatGPT 4o (released May 13, 2024 by OpenAI) in order to turn section outlines into complete initial drafts, and for refining language and improving readability. After using this tool, the authors reviewed and edited the content as needed, and we take full responsibility for the content of the publication.

## Supplemental Information

Document S1. Supplementary Figures 1-3.

Supplementary Table 1. Excel file containing SNOMED concepts used to define endometriosis patients.

Supplementary Table 2. Excel file containing conditions significantly associated with endometriosis at UCSF across the full patient timeline, across thirty control iterations and after filtering through the healthcare utilization-adjusted analysis.

Supplementary Table 3. Excel file containing conditions significantly associated with endometriosis across both the UCSF UCHDW datasets, using the full patient timeline.

Supplementary Table 4. Excel file containing conditions significantly associated with endometriosis at UCSF using only pre-endometriosis diagnoses, across thirty control iterations and after filtering through the healthcare utilization-adjusted analysis.

Supplementary Table 5. Excel file containing conditions significantly associated with endometriosis across both the UCSF UCHDW datasets, using only pre-endometriosis diagnoses.

