## Supplementary material for "Comorbidity analysis and clustering of endometriosis patients using electronic health records": Document S1

**Supplementary Figure 1.** Low-dimensional visualization of endometriosis patients and the closest 1:1 matched controls.

**Supplementary Figure 2.** Volcano plots of all conditions tested for significance.

**Supplementary Figure 3.** Concordance of odds ratios for overlapping significant associations between the analyses performed with the full set of diagnoses and only the pre-endometriosis diagnoses, both at UCSF and UC-wide.

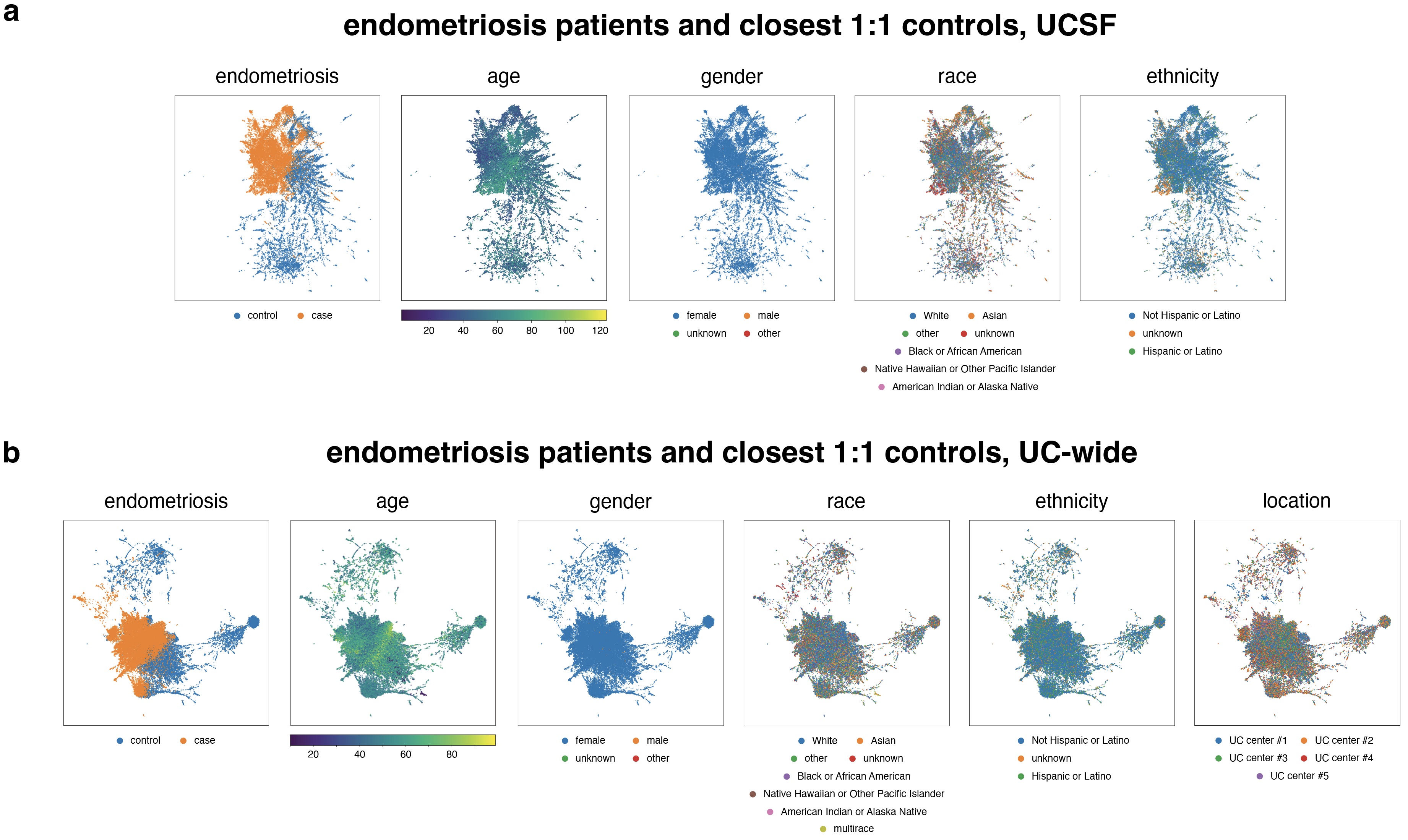

**Supplementary Figure 1.** Low-dimensional visualization of endometriosis patients and the closest 1:1 matched controls. (a) Visualizations of UCSF patients. (b) Visualizations of UC-wide patients.

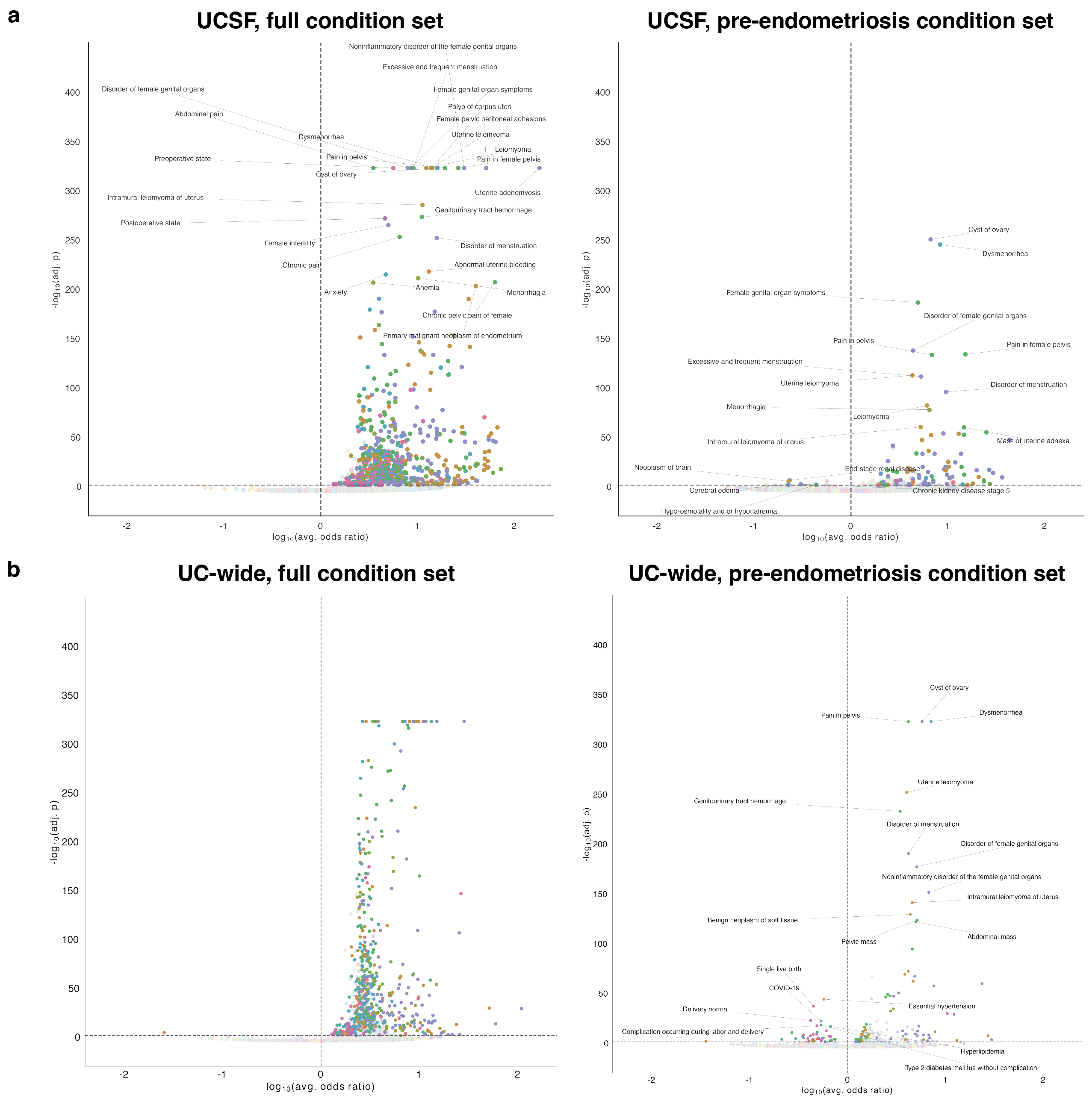

**Supplementary Figure 2.** Volcano plots of all conditions tested for significance. (a) Visualization of UCSF condition sets. (b) Visualization of UC-wide condition sets.

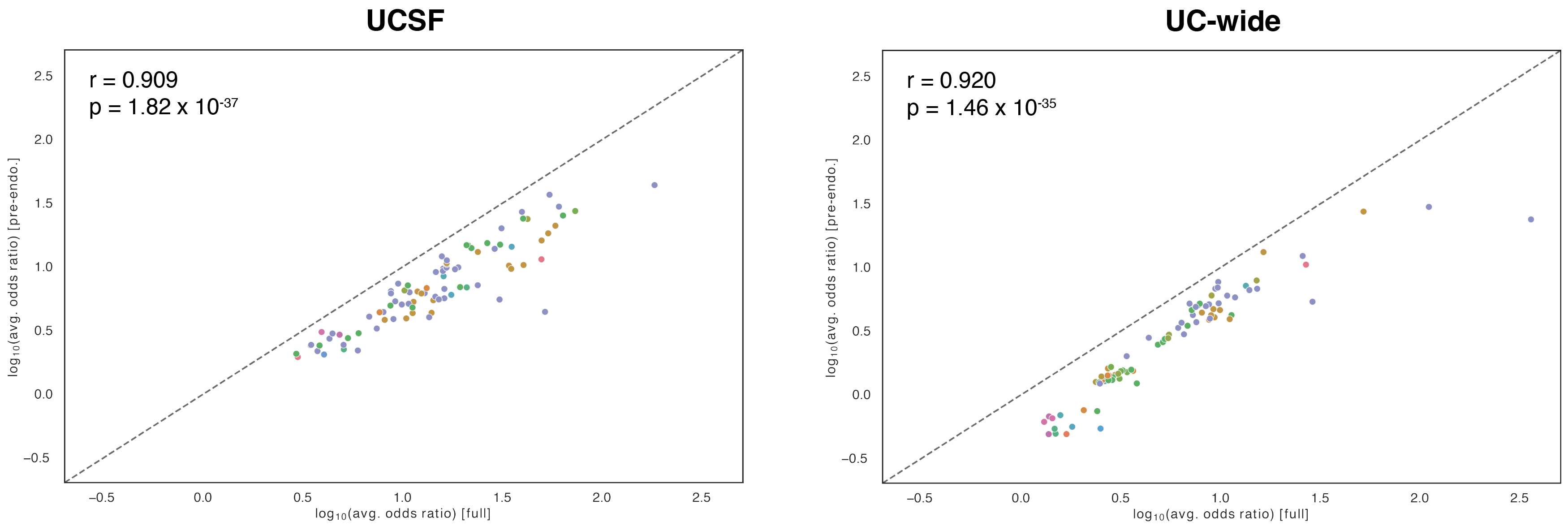

**Supplementary Figure 3.** Concordance of odds ratios for overlapping significant associations between the analyses performed with the full set of diagnoses and only the pre-endometriosis diagnoses, both at UCSF and UC-wide.
